# Ultra-low-field MRI as a tool for measuring brain development in at-risk children in LMICS: feasibility, validity and clinical relevance

**DOI:** 10.64898/2026.06.02.26354785

**Authors:** Layla E Bradford, Jessica E Ringshaw, Thokozile R Malaba, Niall J Bourke, Catherine J Wedderburn, Steven CR Williams, Sean Deoni, Helen Reynolds, Jim Read, Lucy Read, Catriona Waitt, Megan Mrubata, Lee-Ann Stemmet, Lauren Davel, Angela Colbers, Duolao Wang, Saye Khoo, Landon Myer, Kirsten A Donald

## Abstract

**Background:** Children in low- and middle-income countries (LMICs) face an elevated risk of developmental delay, yet scalable neuroimaging tools to study early brain development in these contexts remain limited. Children who are HIV-exposed but uninfected (CHEU) represent a growing population with evidence of language and motor delays and altered brain development compared with children who are HIV-unexposed (CHU). Ultra-low-field (ULF) MRI offers a more affordable alternative to conventional high-field (HF) MRI, but its application in early childhood remains underexplored.

**Methods:** We compared brain volumes derived from ULF (64mT) and HF (3T) MRI in South African CHEU and CHU as part of the DolPHIN-2 PLUS study. Volumetric segmentation was performed using FreeSurfer v7.4.1 and SynthSeg on the Flywheel platform. Agreement between modalities was assessed using Pearson’s and Lin’s concordance correlation coefficients across global and subcortical regions. Associations between ULF-derived brain volumes and developmental outcomes, measured by the Bayley Scales of Infant Development, Third Edition, were evaluated using partial correlations adjusted for sex and age.

**Results:** Forty-five children (9 CHEU, 36 CHU; mean age 45.6 months) had paired ULF and HF scans of usable quality. Strong correlations were observed between ULF and HF volumes for global white and grey matter regions (r > 0.92) and larger subcortical grey matter structures such as the thalamus, caudate, and putamen (r = 0.86–0.89). Moderate-to-weak correlations were evident in smaller structures (hippocampus, pallidum, amygdala). ULF underestimated most grey matter volumes, and overestimated total white matter volume relative to HF. ULF-derived global and subcortical volumes were associated with receptive and expressive communication (r = 0.34–0.59, all p < 0.05).

**Conclusions:** ULF MRI produces brain volume estimates comparable to HF MRI and captures meaningful associations with early language development. These findings support ULF MRI as a feasible and scalable tool for studying neurodevelopment in vulnerable paediatric populations in LMICs.

## Introduction

Neurodevelopmental risk in early childhood is a global public health concern, disproportionately affecting children in Sub-Saharan Africa where infectious diseases, poor nutrition and adverse socio-environmental conditions are prevalent(1). Among the paediatric populations in this region, children who are HIV-exposed but uninfected (CHEU) are emerging as a substantial vulnerable group. With the widespread implementation of programs aimed to reduce vertical HIV transmission, and the introduction of newer and more effective antiretroviral therapy (ART) regimens, a growing number of children are born to mothers living with HIV but do not acquire the virus themselves. Although not infected, CHEU show evidence of language and motor developmental delays, alongside altered brain development in total grey matter and subcortical regions such as the basal ganglia(2-8). As the ART landscape continues to evolve, there is a growing need to measure how in utero HIV and ART exposure influence brain development. The development and validation of contextually feasible tools for measuring brain development in CHEU and other vulnerable paediatric populations in low-and middle-income countries (LMICs) is therefore of critical importance.

Neuroimaging offers unique and valuable insights into the neurobiological basis of development, however access to conventional high-field (HF; ≥1.5T) MRI is currently very limited in LMICs due to high purchase, infrastructural and operational costs. In an effort to address the global disparity in MRI accessibility, there has been a marked shift toward ultra-low-field (ULF; <100mT) MRI systems as a promising alternative, offering greater affordability, allowing for wider use in resource-constrained settings(9, 10). Recent global initiatives such as the UNITY (Ultra Low Field Neuroimaging in the Young); www.unity-mri.com) project have demonstrated the feasibility of implementing ULF MRI at scale in LMICs(10). Initial studies in healthy adults demonstrate strong correlations between HF- and ULF-derived global brain tissue volumes, and one recent study of infants found strong correspondence between HF and ULF-derived volumes for brain regions implicated in antenatal maternal anemia(11, 12). However, ULF performance in assessing brain structure in early life, particularly in smaller subcortical regions, and for identifying developmental risk remains underexplored.

The present study aimed to provide early evidence on the potential of ULF MRI as a tool for measuring brain outcomes in LMICs. Specifically, we assessed the correlation and concordance of brain volume metrics derived from ULF (64mT) HF (3T) MRI in a cohort of South African CHEU and CHU as part of the DolPHIN-2 PLUS study, with a focus on both global and subcortical brain structures previously shown to be altered in the CHEU population. A secondary aim was to explore the potential of ULF MRI to identify developmental risk by assessing the association between ULF-derived brain volumes and developmental outcomes measured using the Bayley Scales of Infant and Toddler Development, Third Edition (BSID-III).

## Methods

### Study Design

This was a cross-sectional sub-study, nested within the DolPHIN-2 PLUS observational follow-up study. CHEU were originally enrolled in the DolPHIN-2 clinical trial (NCT03249181), which evaluated dolutegravir versus efavirenz based antiretroviral therapy (ART) during late pregnancy, while CHU of a similar age were recruited postnatally from the same community(13). Between 2021 and 2023, participants underwent paired ultra-low-field (ULF; 64 mT) and high-field (HF; 3T) T2-weighted MRI scans acquired during natural sleep, alongside developmental assessments using the BSID-III. Children were eligible for inclusion if they had paired ULF and HF scans of usable quality and a BSID-III assessment conducted within six months of imaging. The primary aim was to evaluate the correlation and concordance of brain volume estimates derived from ULF and HF MRI across global and subcortical regions of interest. The secondary objective was to examine associations between ULF-derived brain volumes and developmental outcomes.

### Study Population

Participants were recruited from the Gugulethu Midwife Obstetrics Unit, a primary prenatal healthcare facility located in Gugulethu, an informal peri-urban settlement within the Cape Town metropole with a predominantly isiXhosa-speaking population. Gugulethu, like many other informal settlements in South Africa, is impacted by a significant health burden caused by a high prevalence of HIV. Following informed consent with parents, CHEU previously enrolled in DolPHIN-2 and CHU from the same community were invited to participate when they were between 3 and 4 years of age. Sociodemographic, maternal, and child health characteristics were collected and maternal ART regimen (dolutegravir or efavirenz) and child HIV exposure status were recorded as part of the DolPHIN-2 parent study.

### Measures

#### Sociodemographic characteristics

Sociodemographic, maternal and child health characteristics were collected for all participants. Maternal alcohol exposure during pregnancy was assessed using the Alcohol, Smoking and Substance Involvement Screening Test (ASSIST). Maternal depression was evaluated using the Edinburgh Postnatal Depression Scale (EPDS), with a score of ≥13 indicating risk for depression(14). Maternal employment status and educational attainment were measured using study-created questionnaires. Child HIV exposure status (CHEU vs CHU) and maternal ART regimen during pregnancy (dolutegravir or efavirenz) were determined as part of the parent DolPHIN-2 trial. Study procedures in the pregnant mothers from DolPHIN-2 have been described previously(13, 15)

#### Neuroimaging acquisition and processing

Both ULF and HF MRI were conducted at the Cape Universities Body Imaging Centre during natural, non-sedated sleep using techniques pioneered by our paediatric neurodevelopment research team at the University of Cape Town(16). Scans were acquired between 2021 and 2023.

ULF MRI was performed using the Hyperfine Swoop 64mT system equipped with an 8-channel head coil (software versions 8.2.0 – 8.6.1). Low resolution anisotropic T_2_-weighted images were acquired in three orthogonal planes and reconstructed into a single high-resolution isotropic image (1.5 x 1.5 x 1.5mm^3^) using multi-resolution registration (17). Although T_1_-weighted neuroimaging is conventionally used for morphological assessment, T_2_-weighted neuroimaging was chosen as it demonstrated superior contrast and robustness to motion artefact at ULF. HF MRI was performed using a Siemens Skrya 3T scanner equipped with a 32-channel head coil. High-resolution T_2_-weighted images were acquired using the following parameters: TR = 3200ms; TE = 565 ms; voxel size = 1 x 1 x 1 mm; 176 slices, 1.0 mm thick.

Brain volume segmentation for both ULF and HF scans were performed using the Freesurfer (v7.4.1) recon-all-clinical pipeline built into Flywheel (www.flywheel.io), a centralised and automated medical imaging data storage and processing platform (18). Both global and subcortical volumes were segmented according to the Desikan-Killiany atlas (19). All MRI scans underwent visual quality control (QC) checks by trained researchers to assess motion artefacts, field-of-view coverage, and overall image usability prior to inclusion in volumetric analyses

#### Child Development

Child development was assessed by trained study staff members using the Bayley Scales of Infant and Toddler Development, Third Edition (BSID-III), a standardized tool that evaluates cognitive, language (receptive and expressive), and motor (fine and gross) development(20). BSID is sensitive to subtle developmental delay across cognitive, language and motor scales and has been validated for use in South Africa(20, 21)

### Statistical Analysis

Maternal and child sociodemographic characteristics on all participants with paired ULF and HF data were summarized. Child variables included sex and HIV and ART exposure. Maternal variables included education attainment, employment, alcohol use in pregnancy, and depression.

Descriptive statistics were used to summarize brain volume measurements derived from ULF and HF MRI and included mean and standard deviation. To test for group differences between age at ULF and HF MRI in the same children, paired two-sided t-tests were conducted and also reported. The relationship between ULF and HF-derived brain volumes was evaluated using Pearson’s correlation coefficient (*r*) to assess strength and direction of linear association, and Lin’s concordance correlation coefficient (*P*_*ccc*_) to assess agreement. Regions of interest were chosen a priori based on previous research of CHEU and included total grey matter, white matter, total intracranial volume, and subcortical structures (caudate, putamen, thalamus, pallidum, hippocampus, and amygdala)(6-8). Brain volume outliers were identified using the interquartile range method (brain volume < Q1 – 1.5 × IQR or > Q3 + 1.5 × IQR) and excluded on a region-by-region basis in a sensitivity analysis. To explore the association between neurodevelopmental outcomes and brain volumes, partial correlation analyses were conducted between BSID-III domain scores (cognitive, receptive and expressive language, fine motor, and gross motor domains) and ULF-derived brain volumes in the key regions of interest. These associations were confirmed using HF-derived brain volumes. Analyses were adjusted for child sex and age difference between the ULF scan and developmental assessment.

To account for age-related changes in brain structure, participants were excluded from analyses if the time between their ULF and HF MRI scans, or between the ULF scan and their developmental assessment, was six months or greater. Age outliers were also excluded from the analysis between ULF-derived brain volumes and developmental outcomes. Analyses were conducted using Stata/SE version 18.0 and a two-sided significance threshold for all statistical tests was set at p <0.05. The strength of correlations was interpreted as very strong (*r* ≥ 0.90), strong (0.70 ≤ *r* < 0.90), moderate (0.50 ≤ *r* < 0.70) and weak (0.50 ≤ *r* < 0.70), and concordance as perfect agreement (*P*_*ccc*_ > 0.99), substantial agreement (0.95 < *P*_*ccc*_ ≤ 0.99), moderate agreement (0.90 < *P*_*ccc*_ ≤ 0.950), fair agreement (0.80 < *P*_*ccc*_ ≤ 0.90), and poor agreement (*P*_*ccc*_ ≤ 0.80)(22, 23).

## Results

Sixty-nine participants completed T2-weighted ULF scans, of which 24 were excluded due to either motion and field-of-view artefacts (n = 19) or lack of paired HF scans (n = 5), resulting in 45 participants with paired ULF and HF scans of usable quality (Figure 1). Demographic characteristics are outlined in Table 1. Of the 45 children (58% male) with paired ULF and HF MRI, 9 (20%) were CHEU and 36 (80%) were CHU. Thirteen mothers (29%) completed secondary schooling, 16 (36%) were employed (36%), 20 (44%) reported alcohol use in pregnancy and 9 (20%) screened positive for depressive symptoms (EPDS score ≥ 13).

**Table 1.**
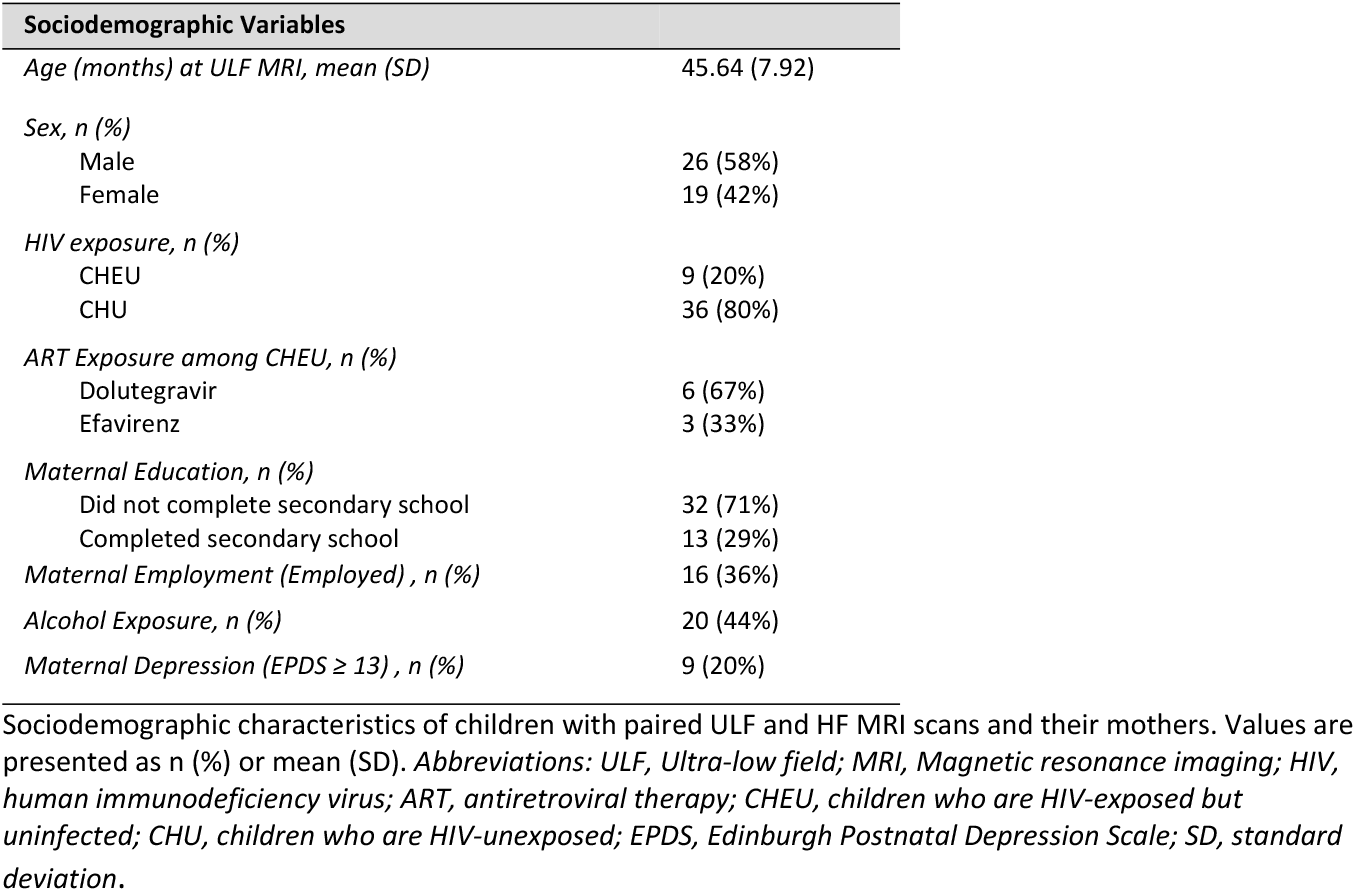
Sociodemographic characteristics of study participants (n=45)

| Sociodemographic Variables |  |
| --- | --- |
| Age (months) at ULF MRI, mean (SD) | 45.64 (7.92) |
| Sex, n (%) |  |
| Male | 26 (58%) |
| Female | 19 (42%) |
| HIV exposure, n (%) |  |
| CHEU | 9 (20%) |
| CHU | 36 (80%) |
| ART Exposure among CHEU, n (%) |  |
| Dolutegravir | 6 (67%) |
| Efavirenz | 3 (33%) |
| Maternal Education, n (%) |  |
| Did not complete secondary school | 32 (71%) |
| Completed secondary school | 13 (29%) |
| Maternal Employment (Employed), n (%) | 16 (36%) |
| Alcohol Exposure, n (%) | 20 (44%) |
| Maternal Depression (EPDS ≥ 13), n (%) | 9 (20%) |

Sociodemographic characteristics of children with paired ULF and HF MRI scans and their mothers. Values are presented as n (%) or mean (SD). Abbreviations: ULF, Ultra-low field; MRI, Magnetic resonance imaging; HIV, human immunodeficiency virus; ART, antiretroviral therapy; CHEU, children who are HIV-exposed but uninfected; CHU, children who are HIV-unexposed; EPDS, Edinburgh Postnatal Depression Scale; SD, standard deviation.

**Figure 1.**
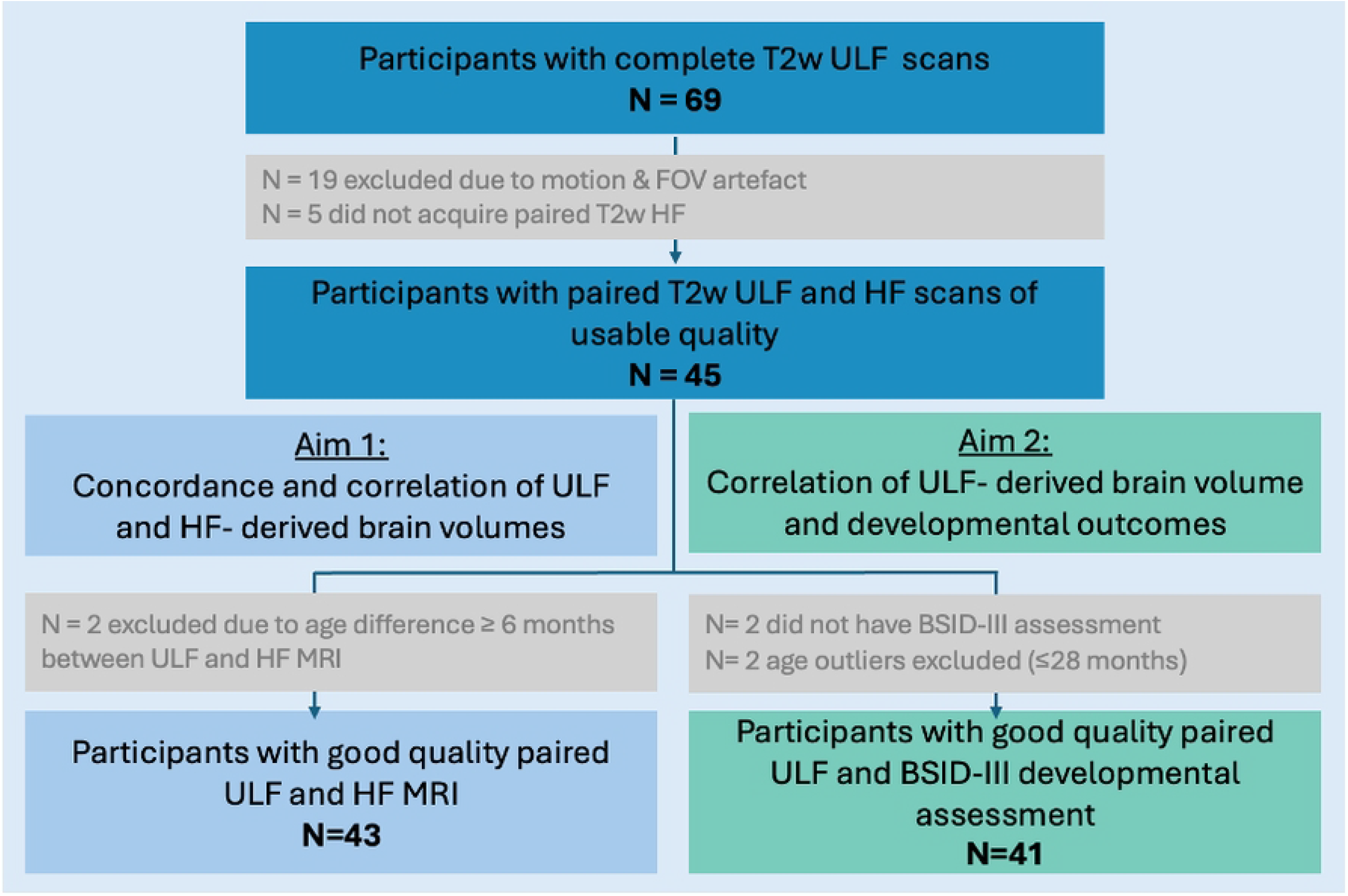
Study flow diagram of participants included in analysis. Flow chart showing participant inclusion and exclusion for the two study aims. A total of 69 children had complete T2-weighted ULF-MRI scans, of which 24 were excluded due to motion/ field of view artefact (n=19) or lack of paired HF MRI (n=5). Of the remaining 45 with usable paired ULF and HF scans, 43 were included in Aim 1 (correlation and concordance of ULF- and HF-derived brain volumes) after exclusion for ≥6 months age difference between scans. For Aim 2 (association of ULF- and HF-derived brain volumes with developmental outcomes), 41 participants were included after exclusion of participants without BSID-III assessments (n=2) and age outliers ≤28 months old (n=2).

### Concordance and correlation of ULF and HF derived brain volumes

In the comparison of ULF- and HF-derived brain volumes, participants were excluded if the interval between ULF and HF MRI was six months or greater (n = 2), resulting in a final sample of 43 included in analyses (Figure 1). Although The 6-month threshold was chosen as the upper boundary, the vast majority (86%) of participants were scanned within 1 month across systems. Strength of agreement between ULF- and HF-derived brain volumes are illustrated in Figure 2. Very strong linear correlations were observed between ULF and HF scans across global brain regions (*r* > 0.92) (Strong linear correlations were observed in total subcortical grey matter volume (*r* = 0.895) as well as the larger individual subcortical structures such as the thalamus (*r* = 0.889), caudate (*r* = 0.881) and putamen (*r* = 0.860). Moderate correlations were found in the pallidum (*r* = 0.633) and hippocampus (*r* = 0.634) and a weak correlation in the amygdala (*r* = 0.417). When assessing concordance, only intracranial volume (*P*_*ccc*_ = 954) reached the threshold for substantial agreement. Fair agreement was found in total white matter volume (*P*_*ccc*_ = 0.841), however poor agreement was found in total cerebral, grey matter and all subcortical structures. As shown in Figure 3, ULF MRI systematically underestimated brain volumes relative to HF MRI across most regions (−0.17 to -15.76% difference), with the exception of white matter (6.15% difference), where ULF values were higher on average resulting in consistently higher volumes. We found no significant differences in age at scan or mean regional volumes between modalities (Supplementary Table S1). Sensitivity analyses excluding brain volume regional outliers did not alter these findings (Supplementary Table S2).

**Figure 2.**
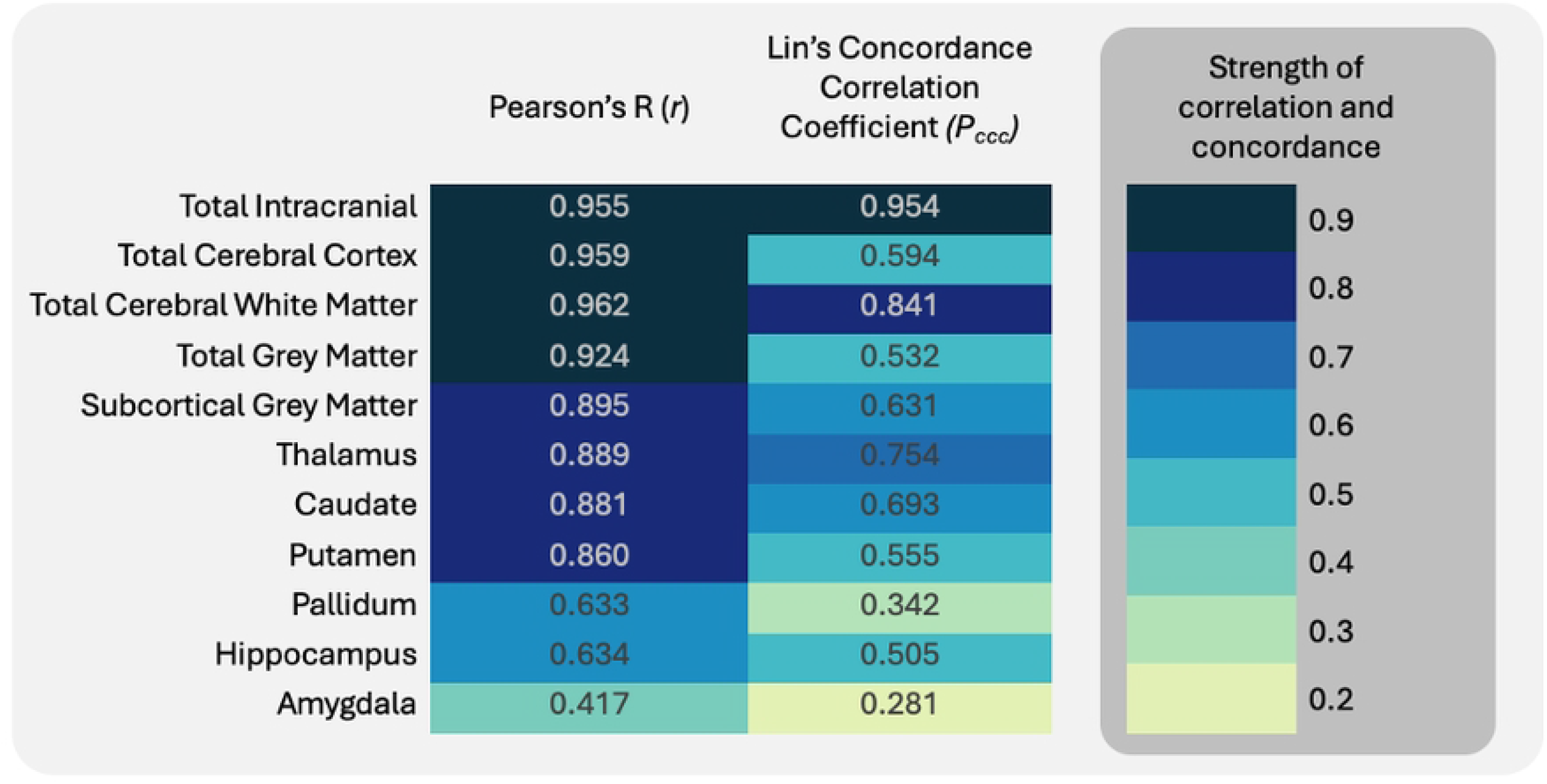
Agreement of brain volume measurements between ultra-low-field (64mT) and high-field (3T) MRI. Heatmap showing Pearson’s correlation coefficients (*r*) and Lin’s concordance correlation coefficients (*P*_*ccc*_) between brain volumes derived from ULF (64mT) and HF (3T) MRI across global and subcortical regions. *Abbreviations: HF, High-field; ULF, Ultra-low field; MRI, Magnetic resonance imaging*.

**Figure 3.**
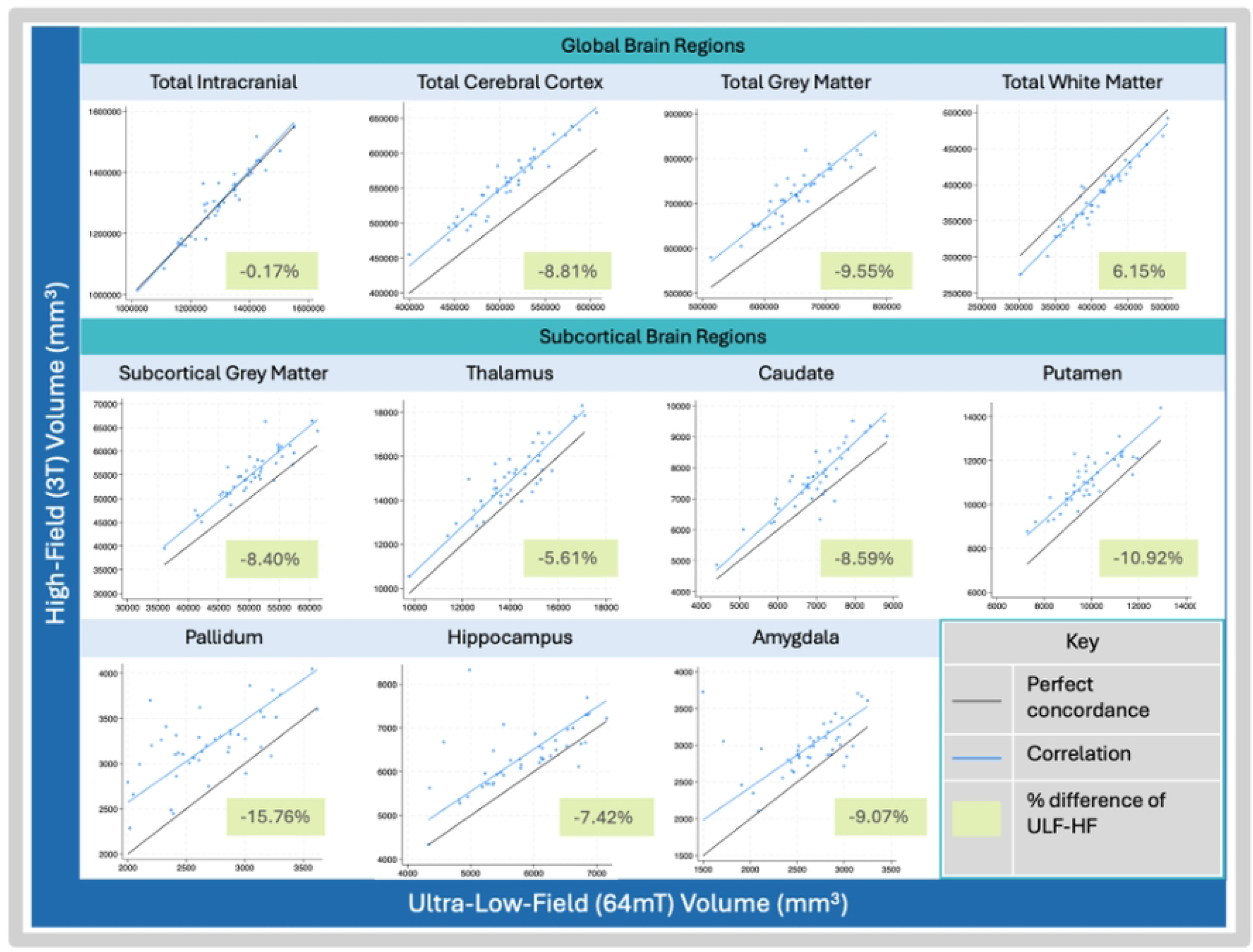
Correlations between ultra-low-field (64mT) and high-field (3T) MRI brain volumes across global and subcortical regions. Scatterplots illustrate the linear correlations (Pearson’s r) between brain volumes derived from ultra-low-field MRI (x-axis) and high-field MRI (y-axis). Line of perfect concordance, correlation and the percentage difference between mean ULF- and HF-derived volumes are presented here. Strong correlations were observed in global brain regions (total intracranial, cerebral cortex, grey matter, and white matter) and several subcortical regions (subcortical grey matter, thalamus, caudate, putamen). *Abbreviations: HF, High-field; ULF, Ultra-low field; MRI, Magnetic resonance imaging*.

### Correlation of ULF-derived brain volume and developmental outcomes

When assessing the associations between ULF-derived brain volumes and developmental outcomes, participants were excluded if they did not have a BSID-III assessment (n = 2) or were age outliers (n = 2; ≤28 months), resulting in 41 participants with both complete MRI and developmental data. Developmental raw scores were calculated for cognitive, receptive and expressive communication, and fine and gross motor domain (Supplementary Table S3). Associations were observed between brain volumes and developmental outcomes (Table 2). At the global level, total grey matter volume was positively associated with receptive (r = 0.37, *p = 0*.*023*) and expressive communication (r = 0.37, *p = 0*.*021*). White matter volume correlated with receptive communication (r = 0.47, *p = 0*.*003*), expressive communication (r = 0.34, *p = 0*.*037*), and fine motor skills (r = 0.35, *p = 0*.*029*). However, the strongest and most consistent associations were observed between ULF-derived subcortical brain volumes and language domains. Subcortical grey matter regions significantly associated with receptive and expressive communication respectively included total subcortical grey matter (receptive communication, r = 0.46, *p = 0*.*004;* expressive communication, r = 0.56, *p < 0*.*001*), thalamus (r = 35, *p = 0*.*029* ; r = 0.42, *p = 0*.*009*), caudate (r = 0.35, *p = 0*.*033* ; r = 0.34, *p = 0*.*036*), putamen (r = 0.44, *p = 0*.*006* ; r = 0.49, *p = 0*.*002*), pallidum, (r = 0.42, *p = 0*.*008*; r = 0.49, *p = 0*.*002*) hippocampus (r = 0.41, *p = 0*.*011*; r = 0.59, *p < 0*.*001*) and amygdala (r = 0.53, *p < 0*.*001*; r = 0.53, *p <0*.*001;* Table 2). The association between ULF-derived brain volumes and communication (receptive and expressive) were confirmed using HF-derived brain volumes for all regions except for the pallidum, where these correlations did not reach significance (Figure 4). All HF-derived brain volumes and development associations are described in Supplementary Table S4.

**Table 2.** Correlations between ULF-derived brain volumes and BSID-III developmental domains.

| Brain Region | BSID-III Domains |  |  |  |  |  |  |  |  |  |
| --- | --- | --- | --- | --- | --- | --- | --- | --- | --- | --- |
|  | Cognition |  | Receptive Communication |  | Expressive Communication |  | Fine Motor |  | Gross Motor |  |
|  | (n=40) |  | (n=40) |  | (n=40) |  | (n=41) |  | (n= 41) |  |
|  | r | p-value | r | p-value | r | p-value | r | p-value | r | p-value |
| *Global* |  |  |  |  |  |  |  |  |  |  |
| Total Grey Matter | 0.281 | 0.087 | **0.369** | 0.023* | **0.373** | 0.021* | 0.303 | 0.061 | 0.284 | 0.080 |
| White Matter | 0.252 | 0.128 | **0.469** | 0.003* | **0.340** | 0.037* | **0.350** | 0.029* | 0.278 | 0.087 |
| *Subcortical* |  |  |  |  |  |  |  |  |  |  |
| Subcortical Grey Matter | **0.373** | 0.021* | **0.462** | 0.004* | **0.558** | 0.000* | **0.399** | 0.012* | **0.338** | 0.035* |
| Thalamus | 0.160 | 0.338 | **0.354** | 0.029* | **0.416** | 0.009* | 0.241 | 0.139 | 0.189 | 0.248 |
| Caudate | 0.311 | 0.058 | **0.346** | 0.033* | **0.341** | 0.036* | 0.241 | 0.139 | 0.279 | 0.086 |
| Putamen | 0.318 | 0.052 | **0.442** | 0.006* | **0.485** | 0.002* | **0.320** | 0.047* | 0.282 | 0.082 |
| Pallidum | **0.450** | 0.005* | **0.423** | 0.008* | **0.494** | 0.002* | **0.443** | 0.005* | **0.384** | 0.016* |
| Hippocampus | **0.463** | 0.004* | **0.407** | 0.011* | **0.593** | <0.001* | **0.470** | 0.003* | 0.309 | 0.056 |
| Amygdala | **0.410** | 0.011* | **0.533** | <0.001* | **0.533** | <0.001* | **0.430** | 0.006* | 0.209 | 0.202 |

Partial correlations (r) between ULF-derived global and subcortical brain volumes and BSID-III developmental domains and p-values are presented here. Correlations adjusted for sex and age difference between ULF MRI and BSID-III developmental assessment. \* $p < 0.05$ . Abbreviations: ULF, Ultra-low field; BSID-III, Bayley Scales of Infant Development, third edition.

**Figure 4:**
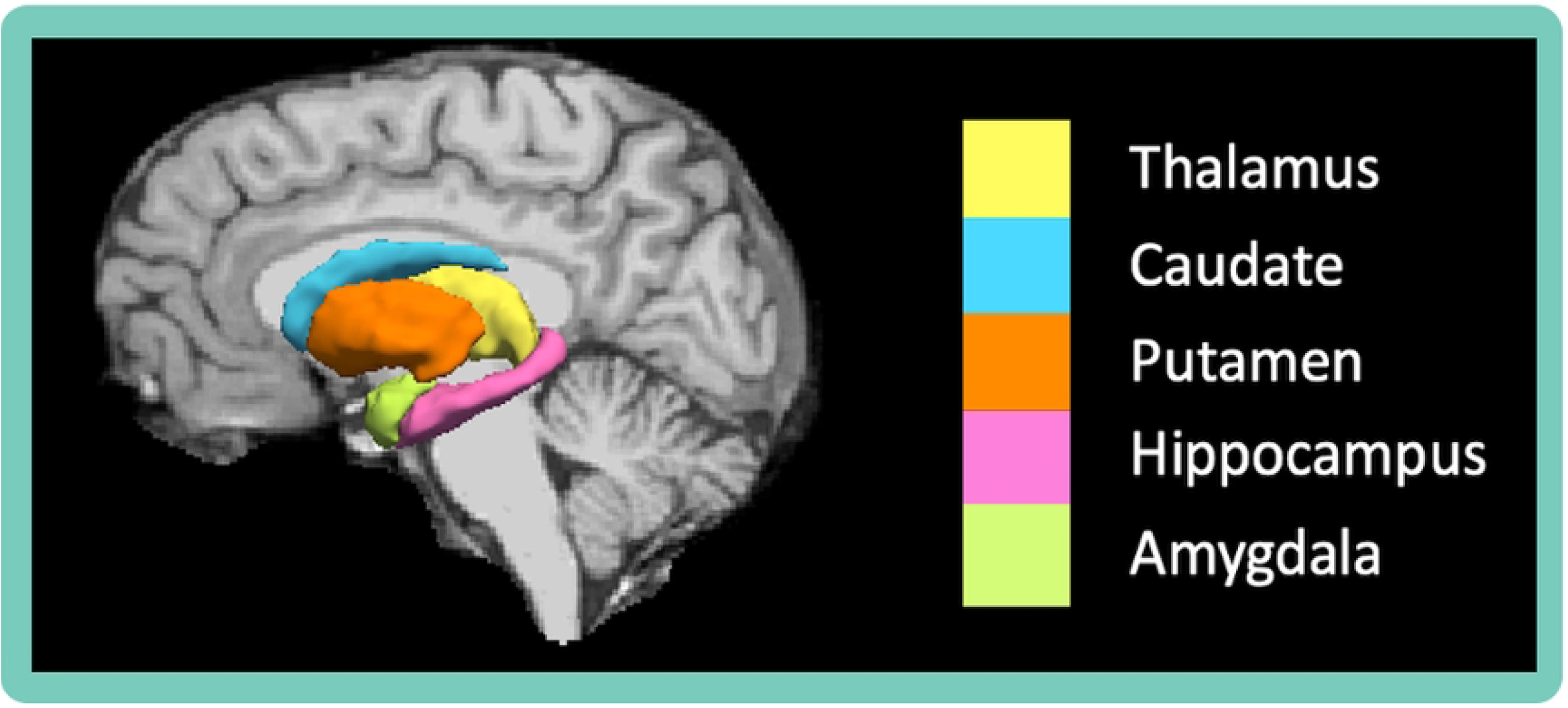
Subcortical regions of interest with significant correlations to receptive and expressive communication across ULF and HF MRI. *Abbreviations: HF, High-field; ULF, Ultra-low field; MRI, Magnetic resonance imaging*.

## Discussion

This study provides early validation of ULF MRI as a feasible and scalable approach to measuring paediatric neurodevelopment in a LMIC setting, with particular relevance for children who are at risk for altered development such as CHEU. Three main findings emerged: first, ULF MRI was feasible to implement in young preschool-aged children; second, ULF produced valid volumetric measurements that correlated strongly with HF MRI; and third, ULF-derived brain volumes demonstrated biologically meaningful associations with developmental outcomes, particularly for language. Although this study does not evaluate the developmental outcomes of CHEU directly, we focused on this population as a clinically relevant example to demonstrate how ULF MRI can be utilized to investigate neurodevelopment in vulnerable populations and highlight this tool as a feasible and scalable approach for use in LMICs.

We successfully acquired 45 paired ULF and HF MRI scans, using non-sedated, natural sleep protocols developed locally to be contextually appropriate in South Africa. The use of natural sleep, rather than sedation, was essential for ensuring both participant safety and the feasibility of implementing ULF MRI in resource-constrained contexts(16). Although 19 ULF scans were excluded due to motion or field-of-view artefacts, this reflects the challenges of one of the first paediatric studies using the ULF Hyperfine Swoop scanner in an African setting. Importantly, these methods have since been refined and expanded through the UNITY network, enabling high-quality ULF data collection at scale across multiple LMIC sites(10, 12).

ULF MRI produced brain volume estimates that strongly correlated with HF-derived volumes for global regions and larger subcortical structures, including the thalamus, caudate, and putamen. Moderate-to-weak correlations were observed in smaller, more complex structures such as the pallidum, hippocampus, and amygdala. This reduced concordance is consistent with the resolution limitations of ULF MRI, where lower spatial resolution increases susceptibility to partial volume effects. These occur when a single voxel encompasses multiple tissue types or adjacent structures, complicating segmentation and volumetric quantification, particularly in small or morphologically complex regions(24). Similar to previous work, ULF systematically underestimated brain volumes relative to HF MRI, except for white matter which was overestimated(25). This pattern suggests grey/white matter boundary definition challenges at low resolution (Figure 5). It also indicates that ULF provides consistent volumetric estimates that could be adjusted or calibrated for use in paediatric neuroimaging. Advances in acquisition software and post-processing pipelines, including image quality transfer (IQT) and super-resolution methods, are expected to further enhance accuracy(17, 26-28).

**Figure 5.**
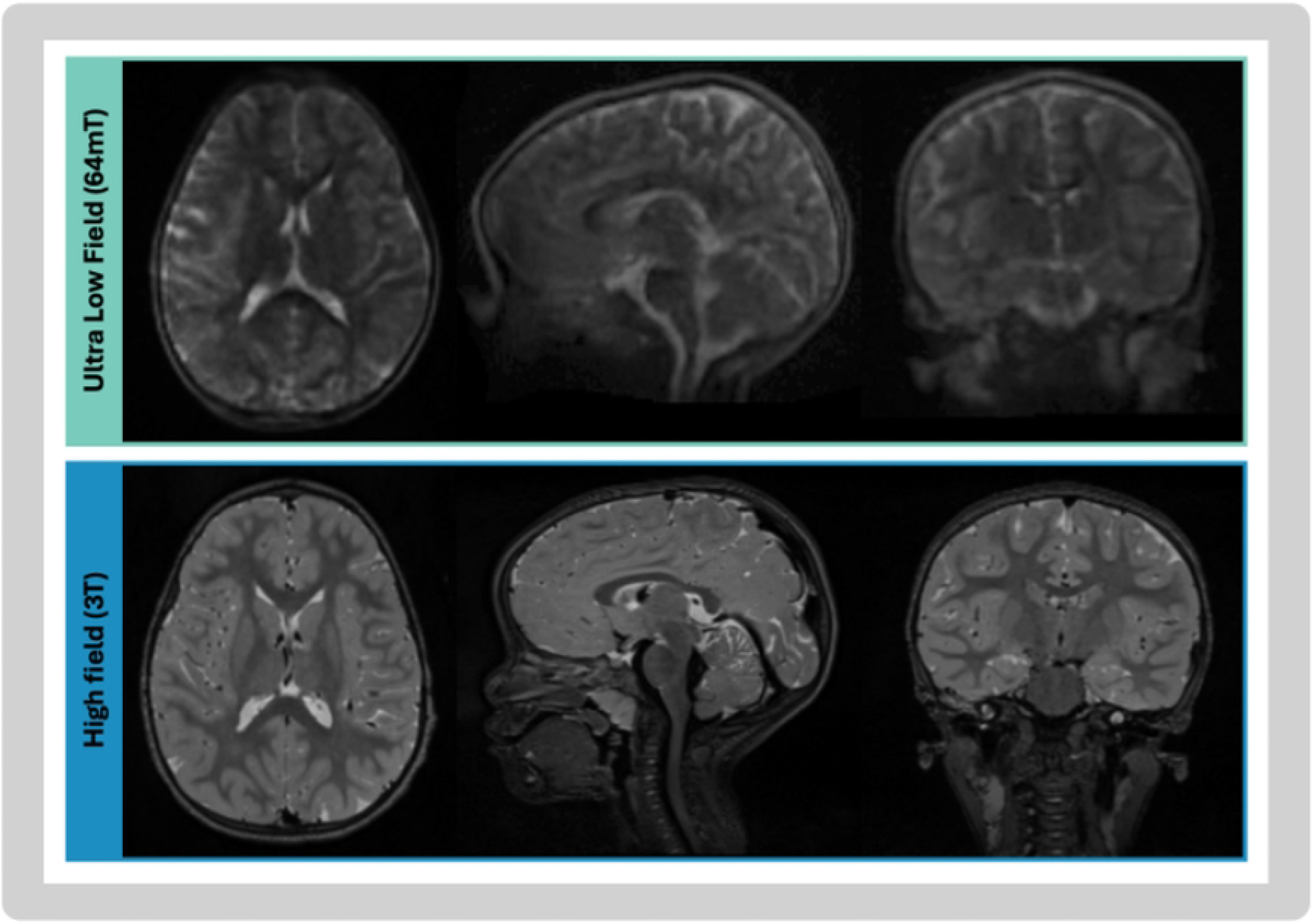
Resolution of ULF and HF T2-weighted MRI scans from children in the DolPHIN-2 PLUS study. *Abbreviations: HF, High-field; ULF, Ultra-low field; MRI, Magnetic resonance imaging*.

We demonstrated associations between brain structure and neurodevelopmental outcomes. The strongest and most consistent relationships were between global (total grey and white matter) as well as subcortical regions of interest (thalamus, caudate, putamen, hippocampus and amygdala) and receptive and expressive communication scales in both ULF and HF (Figure 4). These findings offer clinical significance for the use of ULF MRI in the evaluation of early brain development as communication and language domains have a known vulnerability to early life risk and adversity (29-32). Previous neuroimaging research conducted on HF MRI has similarly shown that global and regional brain volume was a predictor of language outcomes in vulnerable paediatric populations including those with a history of HIV-exposure, preterm birth, neonatal encephalopathy and congenital heart disease (7, 33-36). The agreement of our findings with this prior HF evidence suggests that, despite lower resolution, ULF MRI is sensitive to biologically meaningful brain–behaviour relationships relevant to language development. This strengthens the case for ULF MRI as a feasible and scalable tool to track neurodevelopment in at-risk groups, and may be particularly valuable for improving our understanding of CHEU, who appear especially vulnerable to language delay. The relevance of ULF MRI for researching CHEU extends beyond associations with developmental outcomes. In an era of rapidly evolving ART, careful neurological investigation of ART regimens in CHEU is critical. While recent high-field MRI findings from the DolPHIN-2 PLUS study suggest that newer integrase inhibitor-based regimens such as dolutegravir may not be associated with structural differences in early childhood, the long-term neurodevelopmental implications of in utero ART exposure remain incompletely understood(37). As ART strategies continue to evolve, there is a need for neuroimaging approaches that are feasible in research and clinical contexts to characterise neurodevelopmental trajectories across successive treatment eras. The validation of ULF against high-field MRI in this study therefore provides technical reassurance and supports its potential role in targeted neurodevelopmental research in CHEU as the global ART landscape continues to change.

A key strength of this study is its demonstration of feasibility and validity of ULF MRI in a LMIC context, using locally adapted, non-sedated sleep protocols in a paediatric population. However, there are several limitations to consider. Strict exclusion criteria were applied in order to ensure data quality and comparability, including removal of scans with motion or field-of-view artefacts, age differences greater than six months between paired assessments, and developmental assessments outside the target age window, thus reducing the sample size. The relatively small sample size, while still modest for paediatric MRI studies, limits the generalizability of the findings (11). Additionally, developmental outcomes were assessed using raw BSID-III scores, as many participants were outside of the tool’s normative age range. To address this, we excluded age outliers and adjusted analyses for child age but the use of raw scores may reduce comparability with other similar studies. Lastly, the analyses relied on early versions on ULF acquisition, processing and segmentation software. These pipelines were not originally optimised for ULF images with lower spatial resolution, and therefore reduced accuracy in smaller, more complex subcortical structures. Although steps such as quality control and sensitivity analyses helped mitigate these issues, more advanced processing pipelines such as super-resolution methods and image quality transfer are only now becoming available and may improve future analyses. Ongoing and planned work through the UNITY initiative will expand ULF MRI to multiple LMIC sites, refine acquisition and processing pipelines, and enable longitudinal studies to track developmental trajectories in at-risk populations(10). These efforts have potential to build a foundation for using ULF MRI to inform early identification and intervention strategies for neurodevelopmental risk globally.

## Conclusion

ULF MRI is a feasible and valid alternative to HF MRI. Given its portability and cost-effectiveness, ULF MRI could expand access to neuroimaging in LMICs, enabling better characterisation and understanding of developmental vulnerabilities in at-risk paediatric populations including CHEU.

## Notes

### Ethics Statement

This study was approved by the Faculty of Health Sciences, Human Research Ethics Committee, University of Cape Town (789/2019, 023/2024) and the Central University Research Ethics Committee, University of Liverpool (8730), and written informed consent was obtained during DolPHIN-2 PLUS recruitment.

## Acknowledgements

We thank the DolPHIN-2 PLUS mothers and children for their participation, the contributions of study staff as well as the radiographers at Cape Universities Body Imaging Centre who made this work possible.

## Data Availability Statement

Data cannot be publicly shared due to participant confidentiality and ethical restrictions. De-identified data may be made available upon reasonable request to the corresponding author and with approval from the University of Cape Town Human Research Ethics Committee.

## Supplementary Information

**Supplementary Table S1.** Comparison of paired ultra-low-field (64mT) and high-field (3T) MRI-derived brain volumes.

| Brain Region | ULF (64mT)<br>Mean (SD)<br>N=43 | HF (3T)<br>Mean (SD)<br>N=43 | %<br>Difference | Correlation<br>(*r*) | Concordance<br>(*P<sub>ccc</sub>*) | -value |
| --- | --- | --- | --- | --- | --- | --- |
| Age at scan<br>(months) | 45.74 (7.87) | 46.07 (8.02) |  |  |  | 0.119 |
| *Global* |  |  |  |  |  |  |
| Total Intracranial | 1305312<br>(114581) | 1308745<br>(119235) | -0.17 | 0.955 | 0.954 | <0.001 |
| Total Cerebral<br>Cortex | 505384 (43095) | 554683 (47328) | -8.81 | 0.959 | 0.594 | <0.001 |
| Total Cerebral<br>White Matter | 406057 (41836) | 383553 (43427) | 6.15 | 0.962 | 0.841 | <0.001 |
| Total Grey Matter | 651268 (55617) | 720601 (60372) | -9.55 | 0.924 | 0.532 | <0.001 |
| *Subcortical* |  |  |  |  |  |  |
| Subcortical Grey<br>Matter | 50447.29<br>(5099.85) | 55183.46<br>(5402.934) | -8.40 | 0.895 | 0.631 | <0.001 |
| Thalamus | 14035.01<br>(1478.81) | 14922.44<br>(1529.50) | -5.61 | 0.889 | 0.754 | <0.001 |
| Caudate | 6991.78 (874.33) | 7661.62<br>(1005.77) | -8.59 | 0.881 | 0.693 | <0.001 |
| Putamen | 9892.82<br>(1203.32) | 11113.77<br>(1156.59) | -10.92 | 0.860 | 0.555 | <0.001 |
| Pallidum | 2702.28 (413.54) | 3208.87 (375.15) | -15.76 | 0.633 | 0.342 | <0.001 |
| Hippocampus | 5836.03 (749.42) | 6354.25 (722.32) | -7.42 | 0.634 | 0.505 | <0.001 |
| Amygdala | 2638.69 (387.41) | 2990.62 (342.18) | -9.07 | 0.417 | 0.281 | 0.005 |

Mean (SD) child age, global and subcortical brain volumes at ULF and HF (mean, SD) are presented here (N=43). The percentage difference represents the relative difference of ULF from HF mean volumes. Agreement between paired ULF- and HF-derived brain volumes was assessed using Pearson's correlation coefficients (*r*) and Lin's concordance correlation coefficients (*P<sub>ccc</sub>*). A paired t-test was used to assess differences in age at scan between ULF and HF.
*Abbreviations: HF, High-field; ULF, Ultra-low field; MRI, Magnetic resonance imaging; SD, standard deviation.*

**Supplementary Table S2.** Sensitivity analysis of ULF–HF brain volume correlations and concordance after regional outlier exclusion.

| Brain Region | ULF (64mT)<br>Mean (SD) | HF (3T)<br>Mean (SD) | % Difference | Pearson's R<br>(r) | Concordance<br>(P<sub>ccc</sub>) | P-value |
| --- | --- | --- | --- | --- | --- | --- |
| *Global* |  |  |  |  |  |  |
| Total Intracranial | 1310205<br>(105075.4) | 1312388<br>(109965.1) | -0.17 | 0.945 | 0.944 | <0.001 |
| Total Cerebral Cortex | 504480.9<br>(42599.26) | 553241.4<br>(46982.45) | -9.67 | 0.959 | 0.595 | <0.001 |
| Total Cerebral White Matter | 405521<br>(36258.28) | 382035.9<br>(37211.08) | 5.79 | 0.940 | 0.777 | <0.001 |
| Total Grey Matter | 650429.9<br>(47555.64) | 719120.6<br>(53160.92) | -10.56 | 0.898 | 0.458 | <0.001 |
| *Subcortical* |  |  |  |  |  |  |
| Subcortical Grey Matter | 50747.97<br>(4528.354) | 55404.02<br>(4813.484) | -9.17 | 0.865 | 0.572 | <0.001 |
| Thalamus | 14072.77<br>(1244.092) | 14909.58<br>(1286.83) | -5.95 | 0.823 | 0.672 | <0.001 |
| Caudate | 6993.384<br>(636.9326) | 7650.7<br>(850.7031) | -9.40 | 0.820 | 0.565 | <0.001 |
| Putamen | 9797.312<br>(1098.745) | 10998.25<br>(1039.915) | -12.26 | 0.831 | 0.504 | <0.001 |
| Pallidum | 2711.538<br>(380.3627) | 3218.642<br>(306.9311) | -18.70 | 0.513 | 0.239 | 0.001 |
| Hippocampus | 5899.087<br>(699.7921) | 6371.722<br>(587.9671) | -8.01 | 0.771 | 0.596 | <0.001 |
| Amygdala | 2684.844<br>(259.5096) | 2952.529<br>(220.5819) | -9.97 | 0.599 | 0.362 | <0.001 |

Mean (SD) global and subcortical brain volumes at ULF and HF (mean, SD) are presented here. Statistical outliers (calculated using the interquartile range method) excluded on a region-by-region basis. The percentage difference represents the relative difference of ULF from HF mean volumes. Agreement between paired ULF- and HF-derived brain volumes was assessed using Pearson's correlation coefficients (*r*) and Lin's concordance correlation coefficients (*P<sub>ccc</sub>*). *Abbreviations: HF, High-field; ULF, Ultra-low field; MRI, Magnetic resonance imaging; SD, standard deviation.*

**Supplementary Table S3.** Raw BSID-III developmental assessment scores of study participants.

| BSID-III domain | n | mean | SD | range |
| --- | --- | --- | --- | --- |
| Cognition | 40 | 78.48 | 4.10 | 69 - 86 |
| Receptive Communication | 40 | 40.60 | 4.12 | 30 - 47 |
| Expressive Communication | 40 | 43.58 | 3.46 | 35 - 48 |
| Fine Motor | 41 | 56.24 | 4.84 | 46 - 64 |
| Gross Motor | 41 | 67.83 | 2.64 | 61 - 72 |

Raw scores for BSID-III domains are presented for participants with usable paired ULF and HF MRI scans. *Abbreviations: ULF, Ultra-low field; BSID-III, Bayley Scales of Infant Development, third edition.*

**Supplementary Table S4.** Correlations between HF-derived brain volumes and BSID-III developmental domains.

| Brain Region | BSID-III Domains |
| --- | --- |

|  | Cognition |  | Receptive Communication |  | Expressive Communication |  | Fine Motor |  | Gross Motor |  |
| --- | --- | --- | --- | --- | --- | --- | --- | --- | --- | --- |
|  | (n=40) |  | (n=40) |  | (n=40) |  | (n=41) |  | (n= 41) |  |
|  | r | p-value | r | p-value | r | p-value | r | p-value | r | p-value |
| *Global* |  |  |  |  |  |  |  |  |  |  |
| Total Grey Matter | **0.323** | *0.048\** | **0.479** | *0.002\** | **0.420** | *0.009\** | **0.369** | *0.021\** | **0.345** | *0.032\** |
| White Matter | 0.254 | *0.123* | **0.519** | *0.001\** | **0.387** | *0.017\** | **0.396** | *0.013\** | **0.395** | *0.013\** |
| *Subcortical* |  |  |  |  |  |  |  |  |  |  |
| Subcortical Grey Matter | **0.380** | *0.019\** | **0.520** | *0.001\** | **0.492** | *0.002\** | **0.355** | *0.026\** | **0.380** | *0.017\** |
| Thalamus | 0.290 | *0.077* | **0.422** | *0.008\** | **0.410** | *0.011\** | 0.315 | *0.051* | **0.370** | *0.021\** |
| Caudate | **0.430** | *0.007\** | **0.485** | *0.002\** | **0.432** | *0.007\** | **0.334** | **0.038\*** | 0.312 | *0.053* |
| Putamen | 0.226 | *0.173* | **0.361** | *0.026\** | **0.321** | *0.049\** | 0.179 | *0.275* | 0.225 | *0.169* |
| Pallidum | 0.204 | *0.220* | 0.304 | *0.064* | 0.304 | *0.063* | 0.216 | *0.187* | 0.211 | *0.197* |
| Hippocampus | **0.440** | *0.006\** | **0.556** | *<0.001\** | **0.591** | *<0.001\** | **0.410** | *0.010\** | **0.420** | *0.008\** |
| Amygdala | **0.340** | *0.037\** | **0.520** | *0.001\** | **0.542** | *<0.001\** | **0.326** | *0.043\** | 0.207 | *0.206* |

Partial correlations (r) between HF-derived global and subcortical brain volumes and BSID-III developmental domains and p-values are presented here. Correlations adjusted for sex and age difference between ULF MRI and BSID-III developmental assessment. \* $p < 0.05$ . Highlighted are significant correlations ( $p < 0.05$ ) between brain region and BSID-III domain consistent across both ULF an HF MRI. *Abbreviations: HF, High field; ULF, Ultra-low field; BSID-III, Bayley Scales of Infant Development, 3rd edition.*

